# Clinical cholera surveillance sensitivity in Bangladesh and implications for large-scale disease control

**DOI:** 10.1101/2021.06.02.21258249

**Authors:** Sonia T. Hegde, Elizabeth C. Lee, Ashraful Islam Khan, Stephen A. Lauer, Md. Taufiqul Islam, Taufiqur Rahman Bhuiyan, Justin Lessler, Andrew S. Azman, Firdausi Qadri, Emily S. Gurley

**Author notes:** **Corresponding author:** Dr. Sonia Hegde, Department of Epidemiology, Johns Hopkins Bloomberg School of Public Health, 615 North Wolfe Street Rm E6038, Baltimore, MD 21205. denotes equal contribution (Sonia T. Hegde, Elizabeth C. Lee). **Meetings where information has previously been presented** Asian Conference on Diarrhoeal Disease and Nutrition (ASCODD), January 2020, Dhaka, Bangladesh.

## Abstract

**Introduction:** A surveillance system that is sensitive to detecting high burden areas is critical for achieving widespread disease control. In 2014, Bangladesh established a nationwide, facility-based cholera surveillance system for *Vibrio cholerae* infection. We sought to measure the sensitivity of this surveillance system to detect cases to assess whether cholera elimination targets outlined by the Bangladesh national control plan can be adequately measured.

**Methods:** We overlaid maps of nationally-representative annual *V. cholerae* seroincidence onto maps of the catchment areas of facilities where confirmatory laboratory testing for cholera was conducted, and identified its spatial complement as surveillance greyspots, areas where cases likely occur but go undetected. We assessed surveillance system sensitivity and changes to sensitivity given alternate surveillance site selection strategies.

**Results:** We estimated that 69% of Bangladeshis (111.7 million individuals) live in surveillance greyspots, and that 23% (25.5 million) of these individuals live in areas with the highest *V. cholerae* infection rates.

**Conclusions:** The cholera surveillance system in Bangladesh has the ability to monitor progress towards cholera elimination goals among 31% of the country’s population, which may be insufficient for accurately measuring progress. Increasing surveillance coverage, particularly in the highest risk areas, should be considered.

## Introduction

Bangladesh has among the highest national rates of *Vibrio cholerae* infection in the world [1]; a nationally-representative serosurvey estimated that roughly 17% (95% CI: 11-24%) of the 165 million people living in Bangladesh experienced infection in 2015 [2]. In 2014, the International Centre for Diarrhoeal Disease Research, Bangladesh (icddr,b) and the Institute of Epidemiology Disease Control And Research (IEDCR) established a nationwide sentinel surveillance system with the goal of monitoring the seasonality and geographic trends in acute cases and identifying geographic areas with a high burden of laboratory-confirmed clinical cholera [3]. The participating 22 sentinel hospital sites and the icddr,b Dhaka hospital are the only healthcare facilities that regularly perform laboratory confirmation of *V. cholerae* in the country [3].

The government of Bangladesh proposed their first national cholera control plan in 2019, with the ambitious goals of reducing morbidity and mortality by 50% by 2025 and 90% by 2030 and achieving cholera elimination [4]. Few comparative, nationally representative data exist in Bangladesh, however, with which to compare and measure the reduction in morbidity and mortality. While vaccination campaigns, water, sanitation, and hygiene interventions, and improved case management are the primary tools to achieve these elimination targets, a cholera surveillance system with widespread geographical coverage is necessary to target interventions to the highest burden areas and monitor progress from endemic transmission to elimination.

The US Centers for Disease Control and Prevention (CDC) has a standardized framework for evaluating public health surveillance systems, which may be applied flexibly to systems with varying goals [5,6]. The public health goal of widespread disease control and elimination, like that for cholera in Bangladesh, requires the identification and monitoring of areas with high case counts and high relative risk across the population in a timely manner. However, quantitative evaluations of sensitivity are hard to obtain when the surveillance system is the sole source of data on the occurrence of disease; external data are needed to evaluate the proportion of cases or outbreaks that are identified by the surveillance system [7,8].

Our objective was to determine how the geographic coverage of Bangladesh’s national cholera sentinel surveillance system compares to the distribution of *V. cholerae* infection risk to gauge how well the surveillance system is able to monitor progress towards national cholera control. We also examined how alternate sentinel site selection might improve the sensitivity of the surveillance system to detect high burden areas.

## Methods

### Cholera sentinel surveillance data

There are 23 healthcare facilities known to routinely perform laboratory confirmation of *V. cholerae* among suspected cholera cases in Bangladesh, which includes the 22 sentinel hospital sites in the national cholera surveillance system (in operation since 2016) and the icddr,b Dhaka hospital (Supplementary Table 1). In the absence of specific data on healthcare utilization for cholera at these sites, we assumed that the catchment areas of the different healthcare facility types were as follows: subdistrict (10km), district (20km), tertiary care (30km), and the icddr,b Dhaka hospital (30km) (Figure 1). We refer to the joint set of buffers around all 23 hospitals as the *cholera surveillance zone*, an estimate of the area where suspected cholera cases may be tested and reported.

**Figure 1.**
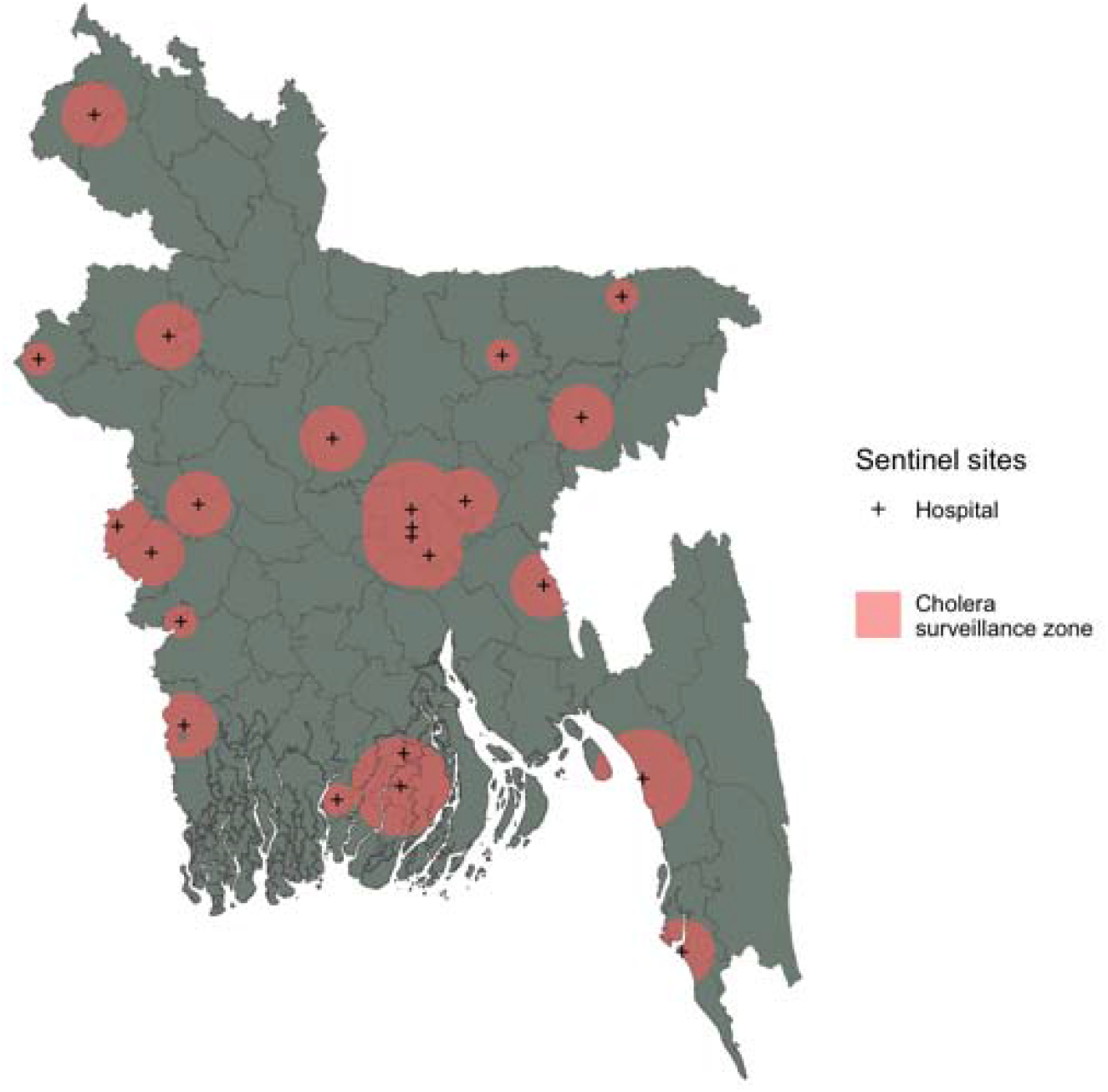
A map of the cholera greyspots in Bangladesh. Populations living in the coral pink areas are inside the cholera surveillance zone. The grey areas are places where we have little information on clinical cases of cholera as they are not captured by the national cholera surveillance system in Bangladesh.

### System sensitivity to detect high cholera case-burden

We used previously published maps of the estimated *V. cholerae* O1 seroincidence from 2015 as the presumed ground truth estimates of burden. These maps include national estimates of the risk of *V. cholerae* O1 infection rates and relative infection risk (compared to the population-weighted mean) across a 5 km x 5 km grid of Bangladesh [2,9] (Supplementary Figure 1A).

We defined the relative and absolute magnitudes of the infection risk and defined thresholds for *high, moderate*, and *low* relative and absolute risk across 25 km^2^ grid cells. We used the 25th and 75th percentiles of the mean grid cell-level risk (relative or absolute) to define cutoffs for moderate and high risk areas.

1. *Relative risk:* This is the seroincidence relative to the population-weighted mean seroincidence across Bangladesh. The moderate and high risk thresholds were 0.81 and 0.91, respectively, where 1 indicates a grid-cell risk equal to the population-weighted mean.
2. *Absolute infections*: This is the estimated number of *V. cholerae* O1 infections in each grid cell in the last year, calculated as the product of the median annual seroincidence and the 2015 population estimates in each grid cell according to WorldPop [10]. The moderate and high risk thresholds were 1,514 and 4,815 infections, respectively.

We define *system sensitivity* with two metrics – the proportion of the population and the proportion of infected individuals living in the cholera surveillance zone. Uncertainty in system sensitivity is reported with the 2.5th and 97.5th percentiles of 1000 posterior draws of the previously published gridded seroincidence estimates [2].

### Examining alternative sentinel site selection with a simulation-based approach

We sought to determine whether system sensitivity could be improved with an alternate selection of sentinel surveillance sites. First, we successfully geocoded 491 of 504 large public hospitals in the Bangladesh Ministry of Health and Family Welfare Facility Registry using the R package tidygeocoder [11] (See details in Supplementary Methods). The geocoded healthcare facilities were then used to examine eight hypothetical strategies of sentinel site selection. The total number of sites (23) and the distribution of facility types (i.e., 4 tertiary care facilities, 12 district-level facilities, and 6 upazila-level facilities) matched that of the current cholera surveillance system across all strategies; we assumed that the system size would remain constant. Only 22 new sites were selected for each set, as the icddr,b cholera hospital in Dhaka remained fixed as a site across all strategies. Twenty simulations (different sets of sites) were drawn per strategy based on a crude estimate of the possible number of unique sets of sentinel sites (491 total facilities/23 sites performing laboratory confirmation). We also estimated the sensitivity of a hypothetical surveillance system that would include all 491 large public hospitals.

One strategy selected the 22 sites randomly from all facilities (*Random*), while another selected sites to match the current allocation of sentinel sites by *Division* (first level administrative units). The remaining six strategies weighted site selection by risk in an attempt to prioritize high-risk areas (See details in Supplementary Methods). For each strategy, we estimated the total population and the annual number of infected people in the proposed cholera surveillance zone. We ran three linear regression models to partition sources of variability for each selection strategy across surveillance zone infections with a random effect across (1000) posterior draws for seroincidence risk and a random effect across (20) simulations in a given strategy. The magnitude of the intraclass correlation coefficient (ICC) for the individual random effect models indicates the relative variability explained by each factor. We reported confidence intervals of system sensitivity as the 2.5th and 97.5th percentiles of the joint distribution of the 20 simulations and 1000 posterior seroincidence draws.

### Code and reproducibility

All analyses were performed in R. Data and source code to reproduce analyses are available at https://https://github.com/HopkinsIDD/bgd_cholera_greyspots with additional details provided in the supplemental appendix. The underlying seroincidence estimates are available at https://github.com/HopkinsIDD/Bangladesh-Cholera-Serosurvey.

## Results

### Surveillance system sensitivity

In the year preceding the 2015/16 serosurvey, 16% (95% CI: 13-23%) or 8 million (95% CI: 4.6-11.9) of the 51 million people living in the cholera surveillance zone had been infected (clinically or subclinically) with *V. cholerae* O1 (Figure 1). The infections occurring in the cholera surveillance zone accounted for 29% (95% CI: 27-35) of the 22.5 million *V. cholerae* O1 infections estimated for Bangladesh during the same period [2].

We estimated that 111.7 million (69%) people in Bangladesh live in surveillance *greyspots*, the geographic area outside of the cholera surveillance zone where suspected cholera is unlikely to be confirmed (Figure 1). The cholera surveillance zone captured 3.4 million (95% CI: 1.8-6.0) annual infected individuals living in high relative risk areas in Bangladesh; 74% (95% CI: 62-78%) of the at-risk population living in high relative risk areas in Bangladesh lived in surveillance greyspots (Table S2). The cholera surveillance zone captured 3.6 million (95% CI: 1.6-6.0) annual infected individuals living in moderate relative risk areas in Bangladesh; 70% (95% CI: 63-73%) of the at-risk population living in moderate relative risk areas in Bangladesh lived in surveillance greyspots. Individuals living in the districts of Rajshahi, Kurigram, and Khulna had high relative infection risk but were unlikely to be captured by the cholera sentinel surveillance system (Figure 2).

**Figure 2.**
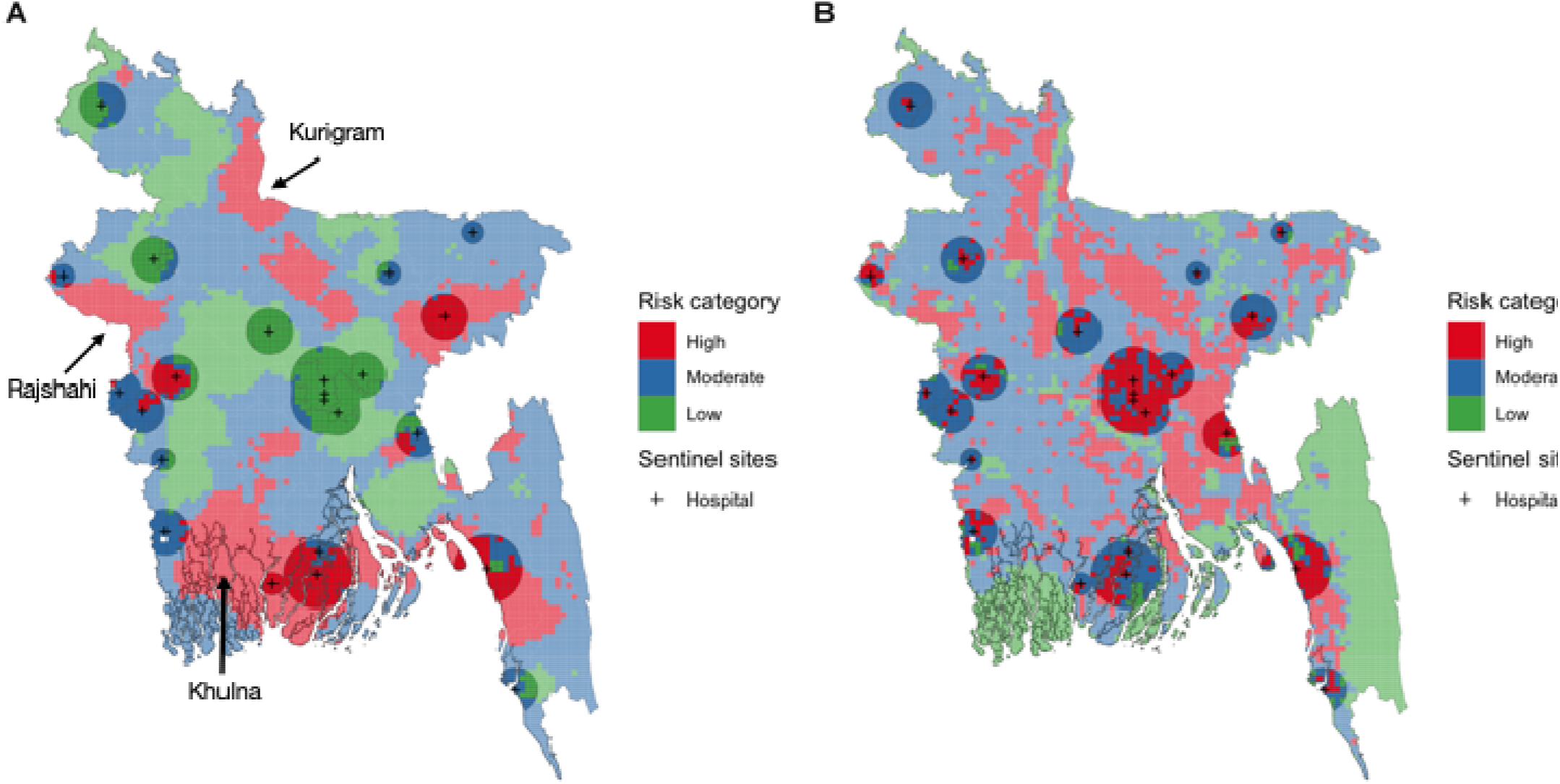
Cholera risk map as categorized by the risk of seroincidence relative to a population-weighted mean by 5 km x 5 km grid cell (Panel A). The map illustrates grid cells of High-Moderate-Low risk and the which grid cells are captured by the cholera surveillance zone (10-20-30-30km for subdistrict, district, and tertiary care, and icddr,b hospitals), indicated by the transparent buffers. Cholera risk map as categorized by the estimated number of *V. cholerae* infections by 5 km x 5 km grid cell (Panel B). The black marks indicate sentinel hospital locations and the transparent buffers overlayed represent the cholera surveillance zone.

Using the absolute risk metric to define risk areas, the cholera surveillance zone captured 6.2 million (95% CI: 3.5-9.2) annual infected individuals living in high absolute risk areas in Bangladesh; 58% (95% CI: 54-59%) of the population living in high absolute risk areas in Bangladesh lived in surveillance greyspots (Table S2). The cholera surveillance zone captured 1.8 million (95% CI: 0.76-2.8) infected individuals living in moderate absolute risk areas in Bangladesh; 78% (95% CI: 74-80%) of the at-risk population living in moderate absolute risk areas in Bangladesh lived in surveillance greyspots.

### Alternative sentinel site selection

Alternative sentinel site selection strategies employing the *Random* or *Division* selection strategy captured a similar percentage of infected individuals in their cholera surveillance zones (16%, 95% CI: 9-23% and 16%, 95% CI: 9-24%, respectively) as the current surveillance system and more directed strategies such as the *Relative Risk Equity* and *Absolute Risk Equity* (18%, 95% CI: 10-25% and 17%, 95% CI: 10-24%, respectively). While percentage differences were small, the mean number of infections captured varied by up to 1 million between some pairs of strategies (Full results in Table S3). If all 491 public facilities were used in the surveillance system, 97% (95% CI: 96.6-97.5%) of infections would be captured (27 million, 95% CI: 16.6 – 37.9).

An examination of the ICC across multiple models of infections in the cholera surveillance zone revealed that there was substantially greater uncertainty in the underlying seroincidence risk estimates than in simulations for the same strategy. The ICC ranged from 0.84 to 0.97 for models with random effects on seroincidence posterior draws, while it ranged only from 0.01 to 0.1 for models with random effects on simulations (full results in Table S4). There were no major differences between strategies.

## Discussion

The cholera sentinel surveillance system in Bangladesh is the only data source available to monitor progress towards national disease control by 2030. Our study described the characteristics of cholera surveillance greyspots, geographic areas where cases are unlikely to be detected because they reside outside the catchment areas of sentinel surveillance sites. We estimated that roughly 111.7 million individuals (69% of Bangladesh’s population) live in greyspots, and that 23% of these individuals (25.5 million people) live in areas with extremely high risk of cholera infection (where the mean annual seroincidence rate is 22%) (Figure 3). The alternative methods for selecting sentinel sites that we explored produced only minor improvements in the capture of cholera infections, although more optimized strategies could be devised. Without changes to the surveillance system, it will be impossible to monitor high cholera burden areas in much of the country, which is a substantive impediment to measuring progress on elimination.

**Figure 3.**
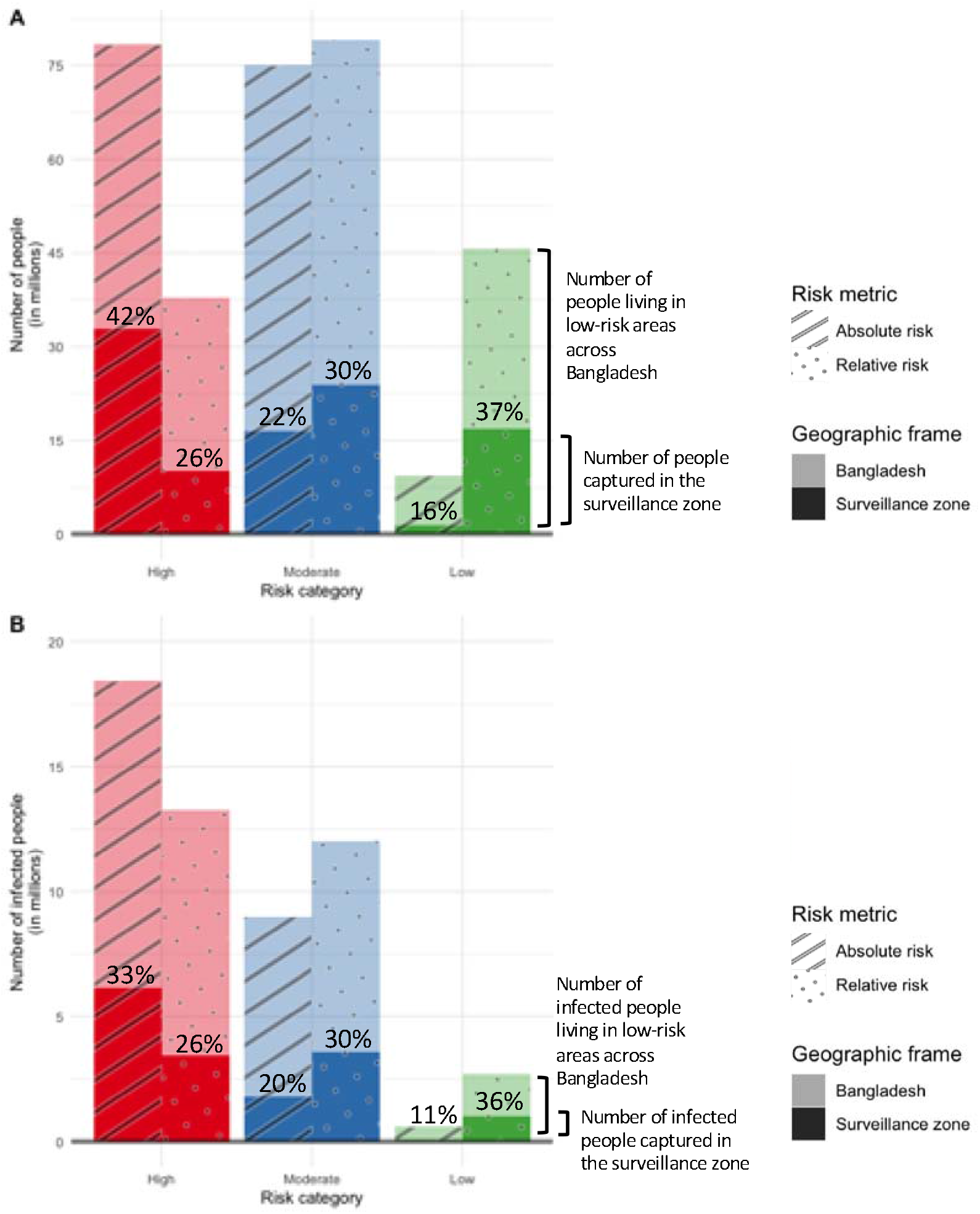
A. The number of people living in high-moderate-low risk areas as defined by the relative and absolute risk metrics across Bangladesh and captured in the cholera surveillance zone (shown in different shades as the geographic frame). The percentages in each bar represent the percentage of the people living in high-moderate-low risk areas across Bangladesh that are captured in the cholera surveillance zone. B. The number of people infected with *V. cholerae* living in high-moderate-low risk areas as defined by the relative and absolute risk metrics across Bangladesh and captured in the cholera surveillance zone (shown in different shades as the geographic frame). The percentages in each bar represent the percentage of infected people living in high-moderate-low risk areas across Bangladesh that are captured in the cholera surveillance zone.

The original stated goals for Bangladesh’s national cholera sentinel surveillance system were to monitor cholera seasonality and epidemiology while also tracking the burden of disease in areas believed to have high prevalence; the objectives may not have had the disease elimination goal in mind. Sentinel surveillance systems that are sensitive to capturing high risk areas are critical to disease elimination efforts to measure disease burden, identify at-risk populations, and monitor the health impacts of interventions in target populations. For cholera control specifically, the drivers of disease transmission are highly local (i.e., fecal-contaminated water and food), with great variation even between households, and campaigns against waterborne diseases must be targeted effectively to high risk areas to achieve success. Depending on whether risk is defined in relative or absolute terms, 58-74% of individuals living in high risk areas were not captured within the cholera surveillance zone. While it is often difficult to quantify the performance of a sentinel system to monitor high risk populations, future work may use our framework to assess how multiple, simulated sentinel selection site strategies may be better suited to achieving different system goals. For example, choosing sites according to population density may best monitor geographic areas with high absolute risk, while choosing sites that are dispersed across geographic divisions may create a sentinel surveillance system that is most representative of national population-level disease trends.

Selecting a strategy to increase overall surveillance sensitivity should consider both cost and feasibility. For example, expanding the number of sites performing laboratory confirmation would expand surveillance system sensitivity, but it may be too costly to be feasible. Furthermore, including all public healthcare facilities in Bangladesh in as sentinel sites would not result in 100% capture of all infections. Future analyses may instead consider the impact of one or more creative solutions, such as the placement of testing sites in select locations that are hard-to-reach and that have high estimated infection risk, or widespread use of cholera rapid diagnostic tests (RDTs). Although RDTs may have lower diagnostic specificity than other laboratory confirmation methods (e.g., 96.5% specificity with Cholkit vs 99.9% with culture) [12], their relatively low cost (2 USD vs 6-8 USD with culture per unit, [13]) and ease of implementation make them prime candidates for expanded use in settings with limited laboratory capacity where the prime purpose for testing is surveillance, not clinical care decisions. Though field evaluations in Bangladesh have demonstrated moderate sensitivity of RDTs relative to culture (e.g., Crystal VC: 72% (95% CI: 51-88%) and Cholkit: 76% (95% CI: 55-91%) [13]), culture tests are known to have a higher false negative rate when antibiotics have been previously taken by the patient and are sensitive to transport conditions when testing is centralized, so sensitivity of RDTs is likely higher than what has been reported [13]. If such tests can be distributed nationally, as stated in the national cholera control plan, the decentralization of testing by use of RDTs may lead to similar if not better performance, real-time tests, and nationwide monitoring for widespread disease control may be feasible when paired with other forms of surveillance.

Our approach has several limitations. We assumed that hospital catchment areas could be defined with simple radial buffers, similar to previous work [14]. A more accurate approach to estimating hospital catchment areas would use patient demographic, symptom, and home address data, and account for barriers to healthcare seeking [15,16]; in reality, the cholera surveillance zone is likely smaller than what we assumed resulting in overestimates of system sensitivity. Conversely, the functional coverage of the cholera surveillance zone may be more expansive than our stated assumptions if private clinics and facilities outside of the national sentinel surveillance system use RDTs or culture to confirm suspected cholera cases, and event-based surveillance systems like media surveillance and hotlines routinely detect disease outbreaks, though samples still have to be processed and confirmed in a lab [17]. Discussions with experts suggest that testing outside of the sentinel surveillance system is low, however, and unlikely to change our results substantially. Finally, while clinical cholera incidence is almost certainly lower than seroincidence, their geographic distributions of burden are likely to be similar and our results should serve as a reasonable proxy for system sensitivity for clinical cholera detection.

The surveillance evaluation framework proposed here, which aims to quantify surveillance system sensitivity to monitoring large-scale reductions in cholera morbidity, may nonetheless prove useful in the context of nationwide control or elimination efforts for other vaccine preventable diseases, like typhoid or Japanese encephalitis [18,19]. By comparing surveillance data to an external validation instrument like a population-representative serosurvey, it is possible to quantify surveillance system sensitivity and perform targeting of interventions that can contribute to an effective elimination strategy. Beyond providing surveillance metrics, an external validation instrument like cross-sectional serology can be used to motivate specific system improvements such as the selection of alternate sentinel sites to increase system sensitivity or even a more cost-effective surveillance system to capture the risk of both asymptomatic and symptomatic infection. Further, by applying multiple definitions of disease risk (e.g., relative versus absolute risk), we can identify surveillance greyspots that are robust to multiple dimensions of information. For example, though the relative risk of *V. cholerae* infection may be considered low in an urban area, the estimated absolute number of infections could be high; we would not want sentinel surveillance sites to be concentrated only in high relative risk areas. Monitoring changes in relative and absolute risk over time, and in rural versus urban areas is important, especially as access to care changes.

Ultimately, the goal of public health surveillance systems are to generate data for action towards improving public health, but if significant gaps in the surveillance system exist such goals may never be met. In Bangladesh, the goal of cholera elimination will likely be hindered by the lack of geographic or population coverage if changes to the system are not made; any documented reductions in morbidity and mortality to quantitate progress will only be among 31% of the country’s population. For any disease, a strong elimination plan should demand high quality surveillance data and using more rigorous and cost-effective methods to evaluate surveillance data is an imperative first step.

## Data Availability

Data and source code to reproduce analyses are available at https://https://github.com/HopkinsIDD/bgd_cholera_greyspots with additional details provided in the supplemental appendix. The underlying seroincidence estimates are available at https://github.com/HopkinsIDD/Bangladesh-Cholera-Serosurvey.

## Acknowledgements

This study was funded by the Bill & Melinda Gates Foundation, National Institutes of Health, and US Centers for Disease Control and Prevention. We thank study staff and participants across Bangladesh for their support.

