## Supplementary Material for "Clinical cholera surveillance sensitivity in Bangladesh and implications for large-scale disease control"

### Supplementary Methods

We filtered public hospitals in the Bangladesh Ministry of Health and Family Welfare Facility Registry of the following types: 200-250 bed, 300-500 bed, general, district, medical college, and upazila health complex [[1]](https://paperpile.com/c/D4qmf1/vSKL). We then used the R package tidygeocoder to obtain geographic coordinates for each hospital using the Google Earth and the Open Street Maps APIs [[2]](https://paperpile.com/c/D4qmf1/u2PV). For each facility, we performed multiple English language searches with the facility name, “Bangladesh,” and different combinations of the division, district, and upazila names. Out of 504 possible hospitals, 491 were successfully geocoded.

One control strategy selected sites randomly from all facilities (Random), while another selected sites to match the number of sentinel sites by first-level administrative units (Division). Beyond the “Random” and “Division” strategies, site selection differed critically in two dimensions: 1) the key indicator used to rank sentinel sites (i.e., population density, mean relative risk of seroincidence in the presumed catchment area, or total absolute seroincidence risk in the presumed catchment area); and 2) the guiding principle behind the spatial distribution of sentinel sites (i.e., “Division” - match the distribution across divisions, “Equity”- at least one site per division before optimizing by site along the key indicator). Twenty-two new sites were selected for each set.

The strategies we compared are as follows:

- Random selection: Select sites randomly. This is a negative control.
- Division selection: Select sites randomly within divisions while matching the current distribution of facilities by division (largest sub-national administrative unit). This is a geographically-stratified negative control.
- Population-Division selection: Within divisions, select sites weighted by population density in their prospective cholera surveillance zones (i.e., Calculate the population density in the 10, 20, and 30 km buffer around upazila, district, and tertiary care facilities, respectively). Match the current distribution of facilities by division.
- Population-Equity selection: There must be at least one site in each of the eight divisions in Bangladesh and these are weighted by population density in the site’s prospective cholera surveillance zone. All remaining sites are selected by population density weight without constraints on division.
- Relative Risk-Division selection: Within divisions, select sites weighted by mean relative risk of seroincidence in their prospective cholera surveillance zones (i.e. Calculate the mean relative risk across all cells in the buffer zone). Match the current distribution of facilities by division.
- Relative Risk-Equity selection: There must be at least one site in each of the eight divisions in Bangladesh and these are weighted by mean relative risk of seroincidence in the site’s prospective cholera surveillance zone. All remaining sites are selected by relative risk weight without constraints on division.
- Absolute Risk-Division selection: Within divisions, select sites weighted by total absolute risk of seroincidence in their prospective cholera surveillance zones (i.e. Calculate the sum of median estimated infections across all cells in the buffer zone). Match the current distribution of facilities by division.
- Absolute Risk-Equity selection: There must be at least one site in each of the eight divisions in Bangladesh and these are weighted by total absolute risk of seroincidence in the site’s prospective cholera surveillance zone. All remaining sites are selected by total absolute risk weight without constraints on division.

### Supplementary Results

**Supplementary Table 1.** Healthcare facilities in Bangladesh that perform laboratory confirmation of *V. cholerae.*

| **Hospital** | **Division** | **Type** |
| --- | --- | --- |
| Upazila Health Complex Bakerganj | Barisal | subdistrict |
| Upazila Health Complex Mathbariya | Barisal | subdistrict |
| General Hospital Patuakhali | Barisal | tertiary |
| District Sadar Hospital Cox's Bazar | Chittagong | district |
| General Hospital Comilla | Chittagong | district |
| Bangladesh Institute of Tropical and Infectious Diseases Chittagong | Chittagong | tertiary |
| District Hospital Norshingdi | Dhaka | district |
| General Hospital Narayanganj | Dhaka | district |
| General Hospital Tangail | Dhaka | district |
| Dhaka Medical College Dhaka | Dhaka | tertiary |
| Uttara Adhunik Medical College Hospital | Dhaka | tertiary |
| District Sadar Hospital Satkhira | Khulna | district |
| General Hospital Kusthia | Khulna | district |
| General Hospital Meherpur | Khulna | district |
| Sadar Hospital Chuadanga | Khulna | district |
| Upazila Health Complex Chaugachha Jesssore | Khulna | subdistrict |
| Upazila Health Complex Madan | Mymensingh | subdistrict |
| Adhunik Sadar Hospital Naogaon | Rajshahi | district |
| Health Complex Shibganj | Rajshahi | subdistrict |
| Adhunik Sadar Hospital Thakurgaon | Rangpur | district |
| Adhunik Sadar Hospital Habiganj | Sylhet | district |
| Upazila Health Complex Chhatak Sunamganj | Sylhet | subdistrict |

**Supplementary Figure 1.** A. Median risk of *V. cholerae* seroincidence relative to a population-weighted mean by 5 km x 5 km grid cell. These relative risk estimates are bounded such that RRs above and below 2 and -2 were plotted as the values 2 and -2, respectively. The black marks indicate sentinel hospital locations. B. Median number of estimated V. cholerae infections per grid cell in the previous year. The black marks indicate sentinel hospital locations. These estimates were calculated as the product of the median seroincidence risk and 2015 WorldPop population estimate in each grid cell. The black marks indicate sentinel hospital locations.

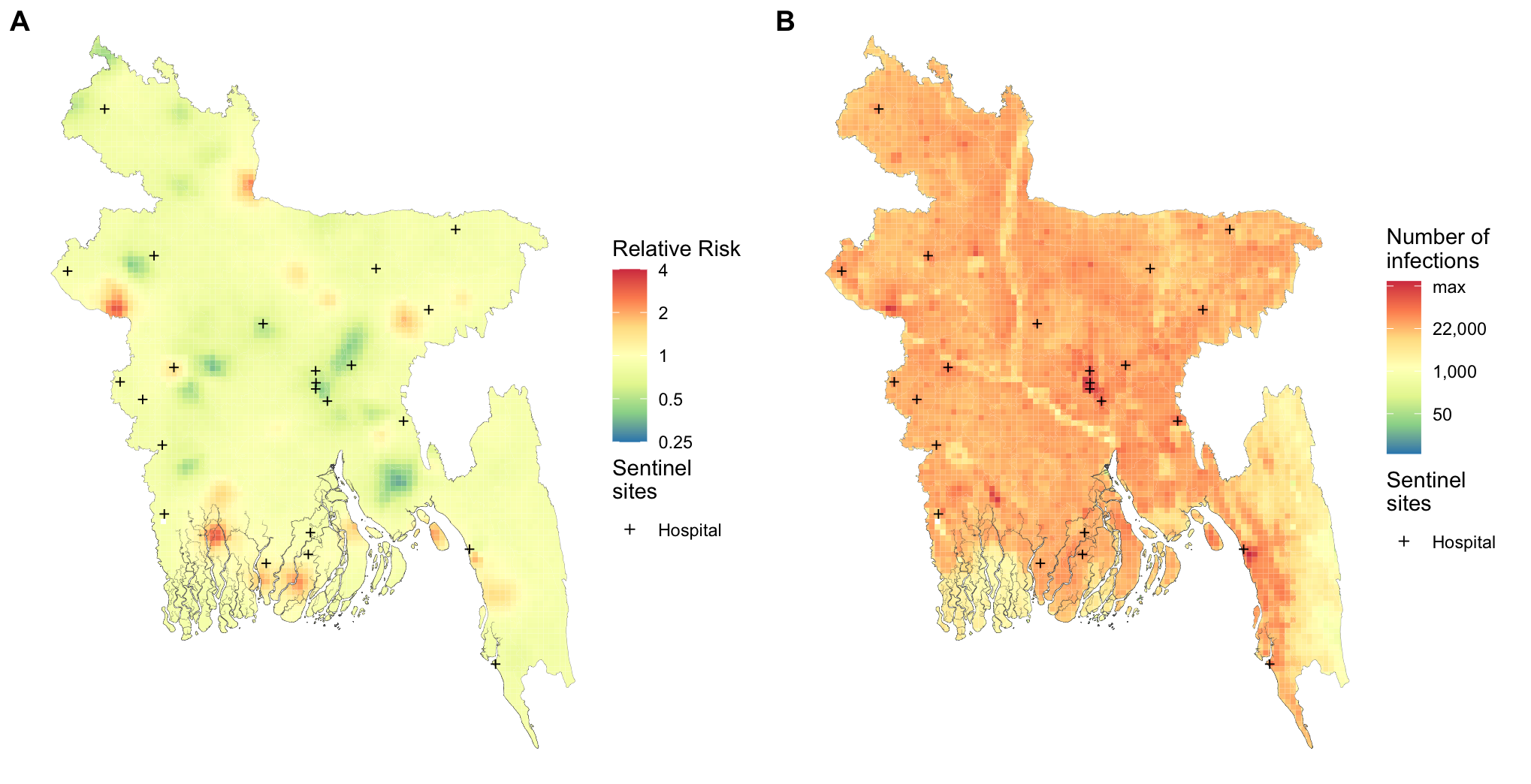

**Supplementary Table 2.** A. The total number and percent of infections that may be captured in the cholera surveillance zone. The percent infected represents the percentage of infected individuals captured within the cholera surveillance zone out of all infected individuals in Bangladesh. B. The number and percent of infections that may be captured in the cholera surveillance zone categorized by the different risk metrics used, relative risk and absolute risk. The infections in the surveillance zone represent the percentage of infected people in high-moderate-low risk grid cells among all infections within the cholera surveillance zone. The population in the surveillance zone represents the percentage of people living in high-moderate-low risk grid cells among all people within the cholera surveillance zone. C. The number and percent of infections that may be captured across Bangladesh categorized by the different risk metrics used, relative risk and absolute risk. The captured at-risk population represents the percentage of high-moderate-low risk populations captured by the cholera surveillance zone out of all high-moderate-low risk populations in Bangladesh. The captured infections represent the percentage of infections in high-moderate-low risk grid cells among all infections in high-moderate-low risk grid cells across Bangladesh.

**A.** *Total cholera surveillance zone population and infections*

| Geographic frame | Population (in millions) | Number infected (in millions) | Percent infected (%) | Percent of all infections in Bangladesh (%) |
| --- | --- | --- | --- | --- |
| Cholera surveillance zone | 50.9 | 8.0 (4.6, 11.9) | 15.7 (13.3 , 23.4) | 28.5 (26.6 , 34.6) |

| **B.** *Population and infections captured in the cholera surveillance zone by risk category* | | | | |
| --- | --- | --- | --- | --- |
| Risk category | Number infected (in millions) | Surveillance zone infections (%) | Surveillance zone population (in millions) | Population in surveillance zone (%) |
| ***Relative risk*** | | | | |
| High | 3.4 (1.8, 6.0) | 43.4 (36.5 , 65) | 10.1 (5.6, 16.5) | 19.8 (16, 32.3) |
| Moderate | 3.6 (1.6, 6.0) | 44.4 (39.4, 58.5) | 23.9 (17.0, 31.5) | 47.0 (41.3, 61.8) |
| Low | 0.99 (0.22, 2.4) | 12.2 (8.0, 26.4) | 16.9 (8.5, 26.3) | 33.3 (26.4, 51.7) |
| ***Absolute risk*** | | | | |
| High | 6.2 (3.5, 9.2) | 77.0 (74.2, 85.9) | 32.8 (27.5, 36.8) | 64.5 (61.6, 72.3) |
| Moderate | 1.8 (0.76, 2.8) | 22.3 (19.6, 30.6) | 16.5 (13.3, 20.4) | 32.5 (29.8, 40.2) |
| Low | 0.062 (0.021, 0.11) | .77 (.6, 1.3) | 1.5 (0.54, 3.6) | 3.0 (1.9, 7.1) |

**C.** *Population and infections captured in Bangladesh by risk category*

| Risk category | Captured at-risk population (%) | Captured infections (%) |
| --- | --- | --- |
| ***Relative risk*** | | |
| High | 26.3 (22.4, 38.1) | 25.9 (21.6, 39.0) |
| Moderate | 30.1 (27.5, 36.7) | 29.7 (27, 36.8) |
| Low | 36.5 (31.9, 49.0) | 35.7 (31.6, 47) |
| ***Absolute risk*** | | |
| High | 41.9 (40.6, 45.8) | 33.3 (30.1, 42.3) |
| Moderate | 22.0 (20.5, 26.5) | 19.8 (19, 22.1) |
| Low | 15.8 (11.5, 27.8) | 10.8 (9.1, 15.5) |

**Supplementary Table 3.** Mean and 95% CI for the number and percentage of the population and infections captured in the cholera surveillance zone across all sentinel site selection strategies.

| Strategy | Millions living in the surveillance zone (95% CI) | Percentage living in the surveillance zone (95% CI) | Millions of infections in the surveillance zone  (95% CI) | Percentage of infections in the surveillance zone (95% CI) |
| --- | --- | --- | --- | --- |
| Random | 53.1 (46.0, 59.8) | 32.7 (28.3, 36.8) | 8.3 (4.5, 12.5) | 15.6 (8.6, 23.1) |
| Division | 56.5 (52.2, 61.9) | 34.7 (32.1, 38.1) | 9.1 (5.0, 13.5) | 16.1 (9.0, 23.6) |
| Population Division | 52.4 (47.7, 58.6) | 32.2 (29.4, 36.1) | 8.5 (4.7, 12.6) | 16.2 (9.1, 23.7) |
| Population Equity | 53.3 (48.3, 61.3) | 32.8 (29.7, 37.7) | 8.2 (4.4, 12.4) | 15.3 (8.4, 22.6) |
| Relative Risk Division | 53.3 (50.0, 55.9) | 32.8 (30.8, 34.4) | 9.3 (5.3, 13.4) | 17.4 (10.1, 25.1) |
| Relative Risk Equity | 50.3 (48.2, 52.0) | 30.9 (29.6, 32.0) | 8.9 (5.2, 12.9) | 17.8 (10.4, 25.5) |
| Absolute Risk Division | 52.4 (49.1, 55.3) | 32.2 (30.2, 34.0) | 8.9 (5.1, 12.9) | 17.0 (9.9, 24.5) |
| Absolute Risk Equity | 55.4 (49.8, 60.9) | 34.1 (30.6, 37.5) | 9.2 (5.3, 13.4) | 16.6 (9.5, 24.0) |

**Supplementary Table 4.** Intracluster correlation coefficients (ICC) for the number of infections in the cholera surveillance zone regressed against different sets of random effects. The purpose of these different models was to partition variability from the underlying seroincidence estimates (1000 posterior draws) and variability from the simulations of each strategy (20 simulations). Three random effects (RE) models were examined: 1) a random effect for each seroincidence posterior draw, 2) a random effect for each simulation in a given strategy, and 3) random effects for both seroincidence posterior draw and simulation.

| Strategy | Seroincidence RE | Simulation RE | Seroincidence and Simulation REs |
| --- | --- | --- | --- |
| Random | 0.84 | 0.10 | 0.94 |
| Division | 0.89 | 0.06 | 0.95 |
| Population Division | 0.92 | 0.05 | 0.97 |
| Population Equity | 0.87 | 0.10 | 0.97 |
| Relative Risk Division | 0.96 | 0.02 | 0.98 |
| Relative Risk Equity | 0.97 | 0.01 | 0.98 |
| Absolute Risk Division | 0.95 | 0.02 | 0.97 |
| Absolute Risk Equity | 0.94 | 0.04 | 0.98 |

**Supplemental Figure 2.** A map of the cholera greyspots in Bangladesh if all 491 possible healthcare facilities from the sentinel site allocation strategy are used to establish the cholera surveillance zone. Populations living in the coral pink areas are inside the cholera surveillance zone. The grey areas are places where we would have little information on clinical cases of cholera in Bangladesh.

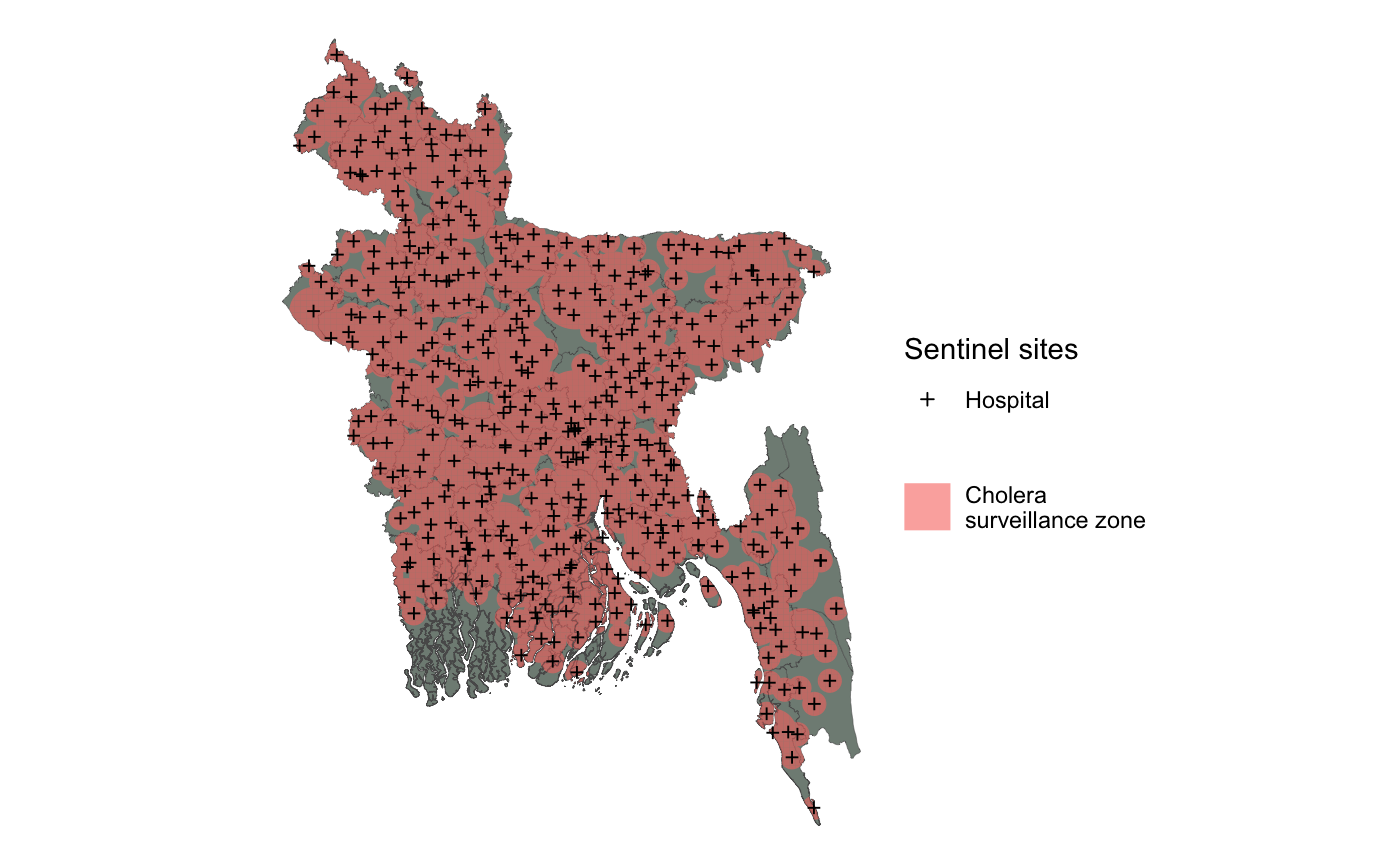
